# An in silico framework for evaluating PRS-guided prognostic enrichment in clinical trial design

**DOI:** 10.64898/2026.03.21.26348974

**Authors:** Ruoyi Cai, Joshua Gillard, Sherry Yang, Lindsay Clegg, Samvel B. Gasparyan, Ying Lu, Andrzej Nowojewski, Lu Tian, Ola Vedin, Euan A. Ashley, Manuel A. Rivas, Jack W. O’Sullivan

## Abstract

**Background:** Clinical trials are essential for therapeutic development but increasingly face challenges due to imprecise inclusion criteria, leading to low event rates and the need for large sample sizes. This inefficiency makes modern trials costly and time-consuming. Despite the availability of extensive clinical, genomic, and biological data, current trial enrollment strategies do not fully leverage this information. Incorporating genomic information into trial design could enable risk-based participant enrichment by preferentially enrolling individuals with higher disease risk, thereby increasing event rates and improving trial efficiency.

**Methods:** In this study, we developed an in silico framework for evaluating prognostic enrichment guided by polygenic risk scores (PRS) in clinical trial design using genomic and electronic health record data from large-scale biobanks. Naturally occurring protective genetic variants were used as analogs of therapeutic interventions, with variant carriers treated as ’treatment’ arms and non-carriers as ’control’ arms. We compared unenriched designs, in which carriers and non-carriers were drawn from the full population, against PRS-enriched designs in which both arms were restricted to participants in the upper 75%, 50%, or 25% of the PRS distribution, respectively. Across these four designs, we quantified disease prevalence, statistical power, sample size requirements, and time-to-event accrual.

**Results:** We applied this approach to the UK Biobank using three model gene–disease pairs: the protective variant p.Arg46Leu in *PCSK9* for coronary artery disease (CAD), p.Gln175His in *ANGPTL7* for glaucoma, and p.Arg381Gln in *IL23R* for inflammatory bowel disease (IBD). Across all three disease contexts, PRS-enriched designs increased disease prevalence, improved empirical power, and accelerated event accrual relative to unenriched cohorts. At 80% power, restricting enrollment to the upper 25% of the PRS distribution reduced required per-arm sample sizes by approximately 60% for CAD-PCSK9 and 78% for IBD-IL23R. Consistent reductions in time-to-event were also observed across enriched strata, suggesting that PRS-enriched trials could achieve target event counts with both smaller sample sizes and shorter follow-up. However, for glaucoma-ANGPTL7, the most restrictive threshold did not yield additional gains over moderate enrichment, as reduced sample size attenuated the detectable difference between arms. These results highlight the need to balance enrichment for higher-risk participants against retaining a sufficient eligible population, and underscore that optimal PRS thresholds are disease-context dependent.

**Conclusions:** These findings establish a generalizable, data-driven framework for prospectively evaluating PRS-guided prognostic enrichment prior to trial initiation. In general, PRS-guided study designs lead to improved empirical power, lower required sample sizes, and faster trials. As population-scale genomic data become increasingly available within healthcare systems and biobanks, this framework provides a scalable foundation for integrating genetic risk information into clinical trial design.

## Introduction

Despite being the gold standard for evaluating therapeutic interventions, traditional randomized controlled trials (RCTs) are often constrained by low event rates, limiting statistical power to detect clinically relevant treatment effects.^1–4^ Currently, only a small fraction of participants may experience the primary endpoint within the study follow-up period. A systematic review of 344 contemporary cardiovascular trials reported a median observed primary endpoint event rate of only 9.0%.^5^ In addition, improvements in background medical therapy and preventive care have further reduced disease risk in the general population.^4,6,7^ Accruing a sufficient number of clinical events in trials typically requires large sample sizes and prolonged follow-up, substantially increasing trial cost and duration.^4,7–9^ Insufficient events lead to null trials, and thus uncertainty whether the medication is beneficial, or if the trial was too small or too short.

To address these challenges, prognostic enrichment strategies have been proposed to improve trial power and efficiency by selectively recruiting individuals with higher disease risk, thereby increasing the expected number of clinical events of interest for a given sample size.^10–15^ However, it remains that current trials use no or few variables to identify and include high risk participants. With the advancement of sequencing technologies, genetic data have become powerful resources for risk prediction at the population level. In particular, polygenic risk scores (PRS) combine the effects of many common genetic variants across the genome to capture an individual’s inherited predisposition to disease.^16^ PRS have demonstrated strong predictive value for a wide range of complex traits and diseases, and can be calculated from birth.^17^ By identifying individuals at higher genetic risk, PRS can enrich trial populations for clinical events, thereby increasing statistical power and reducing the required sample size.^18^

Previous studies have highlighted the potential of PRS to stratify disease risk, enhance clinical risk scores, and have suggested their use for prognostic enrichment in clinical trials.^18–24^ For example, retrospective analyses of completed cardiovascular outcome trials of statins and PCSK9 inhibitors have consistently demonstrated that participants in high-PRS subgroups have a higher event rate and receive a greater relative and absolute clinical benefit.^19,23,24^ A post hoc analysis of the FOURIER trial by Fahed et al. showed that restricting enrollment to individuals in the highest genetic risk strata could theoretically achieve the same level of statistical power with a substantially reduced sample size.^18^ More recently, German et al. incorporated PRS into target trial emulations using observational data and showed that restricting enrollment to participants with higher PRS in emulated cohorts could achieve the required number of events with a smaller sample size than the original trials.^20^

While these studies demonstrate that PRS can meaningfully differentiate risk and identify subgroups with higher event rates, they were retrospective analyses embedded within existing or emulated trials. As such, they evaluated how PRS would have stratified risk within previously conducted trials and were therefore inherently tied to specific completed studies, limiting their applicability for proactively designing enrichment strategies for new trials. Furthermore, prior work generally focused on single diseases within particular trial settings, rather than systematically comparing PRS-based enrichment across multiple gene–disease contexts or alternative risk thresholds. As a result, existing work does not establish a generalizable framework for informing future trial design by quantifying the degree of enrichment achievable under various PRS thresholds through systematic evaluation of sample size requirements, statistical power, and expected event accrual.

To address this gap between the theoretical promise of PRS-guided enrichment and its practical application in prospective trial design, we propose an in silico framework for quantitatively evaluating PRS-based prognostic enrichment using large-scale biobanks with linked genomic and electronic health record data (Figure 1). Our approach leverages naturally occurring protective genetic variants as human genetic analogs of therapeutic inhibition targets, enabling treatment-like effects to be studied without interventional trials. For example, loss-of-function variants in *PCSK9*, such as p.Arg46Leu, lower low-density lipoprotein cholesterol and reduce the risk of coronary artery disease (CAD).^25,26^ Such protective loss-of-function variants have also provided the biological foundation for therapeutic target development, and PCSK9 inhibition is now a well-established clinical strategy.^27,28^ The random distribution of carriers and non-carriers of these naturally occurring protective variants thus creates a “natural experiment,” in which carriers serve as a genetic proxy for the treatment arm and non-carriers for the control arm.

**Figure 1.**
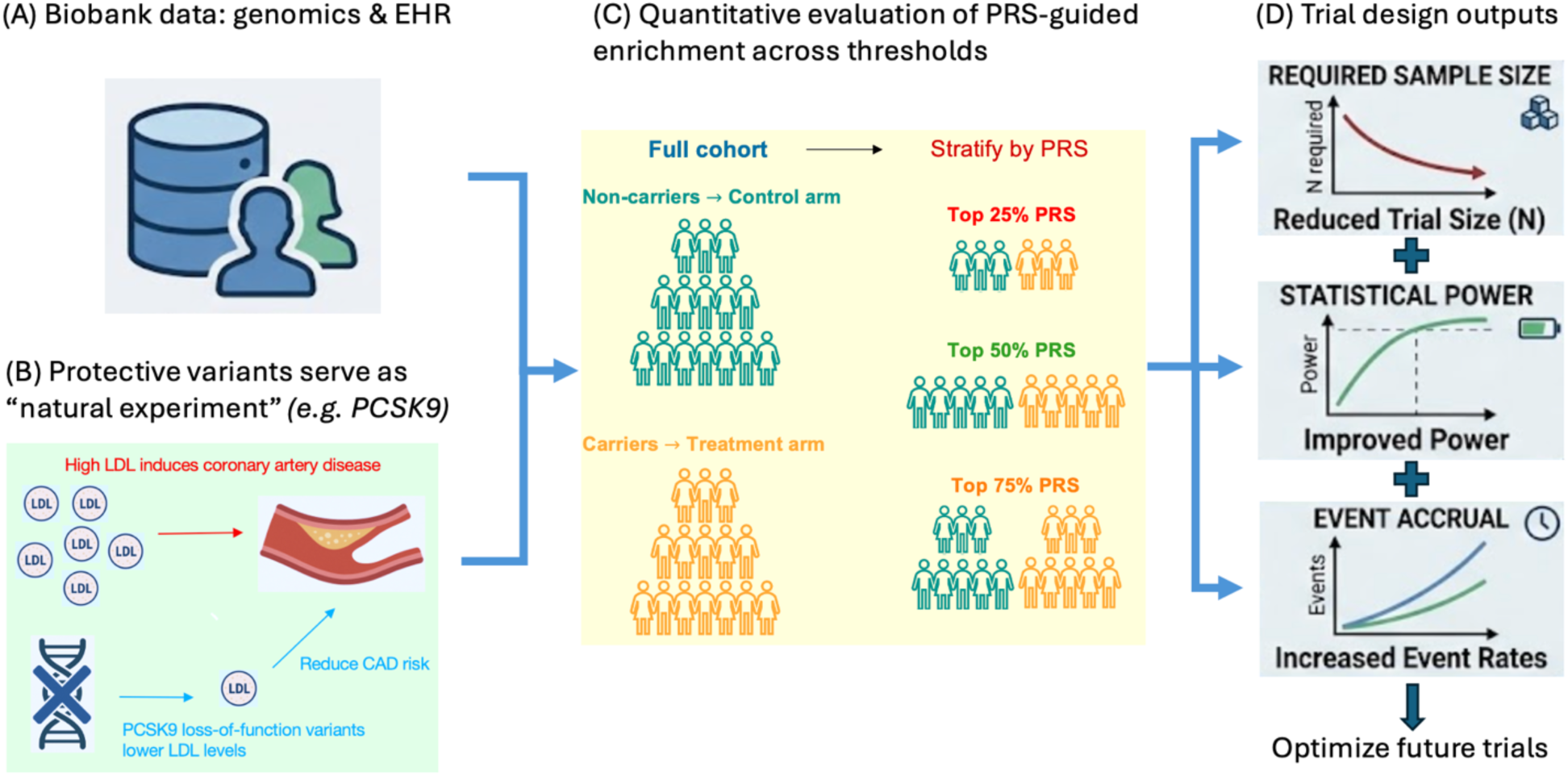
An in silico framework for evaluating PRS-guided prognostic enrichment in clinical trial design using protective genetic variants as therapeutic proxies. (A) Large-scale biobanks with linked genomic and electronic health record data provide the foundation for analysis. (B) Naturally occurring protective variants define a genetic natural experiment in which carriers serve as proxies for therapeutic inhibition and non-carriers serve as controls. (C) Within the full cohort, participants are stratified by polygenic risk score (PRS) thresholds to quantitatively evaluate enrichment across alternative risk cutoffs. (D) Analyses on disease prevalence, required sample size, statistical power, and event accrual within PRS-defined subgroups inform key design parameters for prospective clinical trials.

Using this framework, we stratified participants by PRS quantiles and systematically assessed four key trial design parameters: disease prevalence in carriers versus non-carriers across PRS strata, required sample sizes to achieve target statistical power, empirical power under fixed sample sizes, and prospective event accrual in disease-free cohorts followed over time. Across multiple disease–gene examples, we found that PRS-based enrichment consistently increased disease burden and improved power relative to unenriched designs, although the magnitude of benefit varied by disease context. This approach enables data-driven evaluation of how PRS-based enrichment increases the power to detect treatment effects leveraging protective variants with well-established biological effects, operating entirely using existing population data and feasible prior to trial initiation. Together, these analyses illustrate the potential of human genetics to inform and optimize clinical trial design.

## Methods

### Study population

The study included White British individuals from the UK Biobank.^29^ Individuals were classified as White British based on genetic ethnicity grouping reported in UK Biobank. Participants with missing information on the relevant genetic variants, disease outcomes, or covariates were excluded from downstream analyses.

### Definition of carrier status

Whole-exome sequencing data from the UK Biobank were used to define carrier status for protective genetic variants.^29,30^ For each gene, individuals carrying at least one copy of the protective variant were classified as variant carriers, while non-carriers served as the comparison group. Within the in silico framework, carriers and non-carriers were treated as genetically defined “treatment” and “control” arms, respectively, reflecting the treatment-like effects of these biologically validated variants.

### Definition of disease endpoints

Disease phenotypes were derived from linked hospital inpatient electronic health record data in the UK Biobank and defined using International Classification of Diseases, 10th Revision (ICD-10) diagnosis codes recorded in primary or secondary diagnostic positions.^29^ The ICD-10 code definitions of all disease endpoints considered in this study were summarized in Table S1. Disease status was defined as a binary indicator reflecting the presence of at least one relevant ICD-10 diagnosis code recorded during UK Biobank follow-up.

### Polygenic risk score

Disease-specific polygenic risk scores (PRS) were obtained from the UK Biobank PRS released by Thompson et al. (2022).^31,32^ Within the White British study population, we defined three PRS strata, “Top 75%”, “Top 50%”, and “Top 25%” using the 25th, 50th, and 75th percentiles of each disease-specific PRS, respectively. Individuals above each percentile threshold *q* were assigned to the corresponding “Top (1 − *q*)%” PRS group, and the full study population, labeled as the “All” group, served as the reference group for comparisons with the enriched groups.

### Selection of disease-gene examples for implementing the PRS-guided in silico framework

The workflow used to identify disease–gene examples appropriate for implementing the PRS-guided prognostic enrichment is summarized in Figure S1. We began with a structured literature search to identify candidate disease–gene pairs. Eligible pairs were required to involve well-characterized monogenic variants demonstrating protective effects against the disease of interest, supported by prior genetic or functional evidence.

For each disease–gene pair with verified biological foundation, we evaluated three empirical features to determine whether sufficient statistical information is available in the finite dataset to evaluate PRS-guided enrichment framework. The first feature was the presence of a genetic loss of function variant in the source population, assessed by estimating the odds ratio comparing carriers to non-carriers leading to a significant difference in disease occurrence. This was quantified by examining disease occurrence between carriers (of the variant) vs non-carriers via the Wald statistic and corresponding p-value. Variants demonstrating a statistically significant difference (using nominal level of 0.05) in disease risk between carriers and non-carriers in the full cohort were considered for further evaluation in our PRS-guided trial design framework.

The second feature was whether the PRS meaningfully stratified disease burden. This was evaluated using the odds ratio per standard deviation of PRS and by examining disease prevalence across progressively enriched PRS strata. A statistically significant association between PRS and disease, accompanied by a generally increasing pattern of disease prevalence across strata, indicates that restriction to higher PRS groups shifts disease risk in the intended direction.

The third feature assessed whether the difference in disease risk between carriers and non-carriers remained statistically significant within PRS-enriched strata. Specifically, we evaluated whether the association observed in the overall population persisted when restricting to individuals above the 25%, 50%, and 75% thresholds of the PRS distribution using the stratum-specific carrier odds ratio, its Wald statistic, and corresponding p-value. Because restriction to higher PRS strata reduces the effective sample size and the number of carrier–case observations, the variance of the carrier effect estimate may increase substantially after restriction, and the carrier effect may no longer reach statistical significance in enriched subgroups even if it is significant in the full cohort. This typically reflects insufficient sample sizes of enriched PRS strata and suggests that overly aggressive restriction to high PRS strata is unlikely to yield meaningful efficiency gains.

We summarize the three empirical features described above using a joint information score, denoted by *Ψ*, to provide a quantitative summary of the empirical support for applying the PRS-guided enrichment framework to each candidate disease–gene pair:

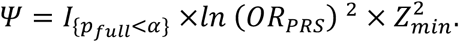

This metric combines three components, corresponding to the presence of a carrier-non-carrier difference in disease prevalence, the magnitude of polygenic risk stratification, and the strength of the carrier association within enriched PRS strata, respectively. The first component, 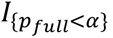, is an indicator function equal to one when the p-value associated with the carrier effect in the full cohort (*p*_*full*_) satisfies the nominal significance threshold *α*, and zero otherwise. The second component, *In* (*OR*_*PRS*_) ^2^, represents the squared log-odds ratio per standard deviation of PRS estimated in the full cohort, capturing the magnitude of the association between PRS and disease prevalence. In the third term, *Z*_*min*_ is the minimum Wald Z-statistic for testing the carrier effect across the evaluated PRS-enriched strata, which reflects the smallest stratum-specific signal to noise ratio for the carrier effect under increasing PRS restriction.

We note that these features focus on whether the available sample size and event counts allow stable estimation of the protective variant effect under PRS restriction, rather than on therapeutic importance or magnitude of biological effects. The latter considerations mainly relate to the operational implications and translational impact but do not determine whether the framework can be assessed with adequate statistical power in the observed data.

### Evaluation of PRS-guided prognostic enrichment

We first examine disease prevalence among protective carriers and non-carriers across PRS-defined subgroups (top 25th, 50th, 75th). Within each PRS-defined stratum, disease prevalence was estimated separately for carriers and non-carriers, and compared.

We next estimated sample size requirements and statistical power based on disease prevalence across PRS-enriched groups to quantify how enrichment by PRS influences trial design parameters. To estimate disease prevalence in each arm for sample size calculations, we fit a logistic regression model of disease outcome on PRS, carrier status of the protective variant, and their interaction (PRS × carrier), adjusting for age, sex, and the first three genetic principal components. For each observed individual, the carrier disease probability was estimated from the fitted logistic regression model assuming the individual is a carrier, while maintaining their observed PRS, age, sex and genetic principal components unchanged. Then for each PRS-defined subgroup, we obtained the average carrier disease probabilities across individuals in the subgroup as the average carrier disease prevalence for the subgroup. Similarly, we calculated the average non-carrier disease prevalence for each PRS-defined subgroup. These population-level disease prevalences were then used for sample size calculation in PRS-based enrichment study design, whose target population is a PRS-defined subgroup.

Sample sizes were calculated based on standard two-proportions Z test at a two-sided 5% significance level across power targets ranging from 60% to 85%. Specifically, the required sample size per group was given by

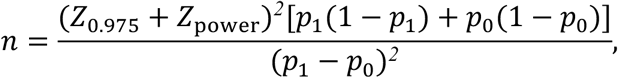

where *Z*_*α*_ is the *α*-th quantile of a standard normal, and *p_1_* and *p_0_* are carrier and non-carrier disease prevalences for the PRS-defined subgroup, respectively. To further assess the recruitment feasibility and operational implications of PRS-based enrichment, we additionally estimated the required screening sample size and trial cost using a streamlined cost framework incorporating participant screening, PRS ascertainment, and trial execution costs based on the calculated sample size. Details of the screening and cost assumptions and calculations are provided in the Supplementary Methods.

Finally, we conducted simulations to empirically quantify statistical power achievable under different combinations of sample size and PRS-based enrichment. Within each PRS-defined subgroup, we constructed synthetic datasets with 100, 200, 500, and 1000 individuals per arm (carriers vs non-carriers) by sampling the corresponding number of individuals with replacement from carriers and non-carriers, separately. Each generated dataset was analyzed using logistic regression with disease status as the binary outcome and carrier status as the independent variable of the primary interest adjusting for age, sex, and three genetic principal components. This process was repeated 1,000 times for each given combination of sample size and PRS-based enrichment, and the empirical power was calculated as the proportion of replicates in which the carrier effect reached nominal statistical significance. To be conservative, when fewer than five disease cases were observed in either group within a resampled cohort, we recorded the comparison as a non-rejection without formally testing the carrier effect to avoid unstable estimates arising from extremely small numbers of events.

### Simulation of prospective event accumulation under PRS-guided enrichment

The PRS-based enrichment can not only increase the overall disease prevalence but also reduce the length of follow-up time to observe a sufficient number of disease events. To quantify how PRS-guided enrichment influences the waiting time to incidence disease, we further analyzed individuals who were free of the relevant disease at baseline, defined as the date of UK Biobank enrollment. Participants were followed prospectively from baseline until incident disease or censoring. Incident disease was defined as the first recorded diagnosis after baseline, and censoring was defined at 10 years of follow-up.

Restricted mean survival time (RMST), defined as the area under the survival curve up to a prespecified truncation time, was used to summarize average event-free time over the follow-up horizon. Within each PRS-enriched subgroup and for each specific target sample size, we generated a synthetic dataset by sampling the corresponding number of individuals with replacement from the baseline disease-free cohort. Then, the RMST up to 10-years was estimated based on the synthetic data. This process was repeated 1,000 times and the empirical distribution of estimated RMST was obtained. We then compared the distribution of RMST across PRS-enriched groups to evaluate how PRS-based recruitment influences the rate of prospective event accrual.

## Results

### Selection of candidate disease-gene pairs

Through our structured literature review, we first identified loss of function variants that confer natural protection against disease. The identified examples included *PCSK9* p.Arg46Leu for coronary artery disease (CAD-PCSK9),^25–28^ *ANGPTL7* p.Gln175His for glaucoma (glaucoma-ANGPTL7),^33–36^ *IL23R* p.Arg381Gln for inflammatory bowel disease (IBD-IL23R),^37–40^ *IL6R* p.Asp358Ala for rheumatoid arthritis (RA-IL6R),^41–44^ *SLC30A8* p.Arg138Ter for type 2 diabetes (T2D-SLC30A8),^45–48^ and *APOC3* p.Ala43Thr for coronary artery disease (CAD-APOC3).^49–52^

We then calculated the joint information score for each disease-gene pair to determine if it met our criteria for inclusion in our framework. To do this, first we obtained GRCh38 genomic coordinates for the protective variant in each disease-gene pair, as well as the ICD-10 code definition of the corresponding disease endpoint are summarized in Table S1. Second, we used the UK Biobank PRS release to obtain PRS for coronary artery disease, glaucoma, type 2 diabetes, and rheumatoid arthritis. Because inflammatory bowel disease was defined in our study as a composite endpoint including Crohn’s disease and ulcerative colitis, and no unified IBD PRS was available, we constructed an IBD PRS proxy by taking the maximum of the CD PRS and UC PRS for each individual. This definition aligns the genetic risk stratification with the composite disease endpoint and prioritizes sensitivity to genetic risk for either IBD subtype.

These data allowed us to calculate joint information scores for each disease-gene pair, which are presented in Figure 2 and detailed in Table S2. Three disease–gene pairs meet our criteria for inclusion (score > 0) and were selected for full in silico enrichment analysis: CAD-PCSK9, glaucoma-ANGPTL7, and IBD-IL23R. PCSK9-CAD and IL23R-IBD satisfied all selection criteria. In the full cohort, both showed a statistically significant odds ratio comparing carriers with non-carriers, as well as a strong association between PRS and disease risk. The carrier versus non-carrier odds ratio remained detectable across progressively enriched PRS strata.

**Figure 2.**
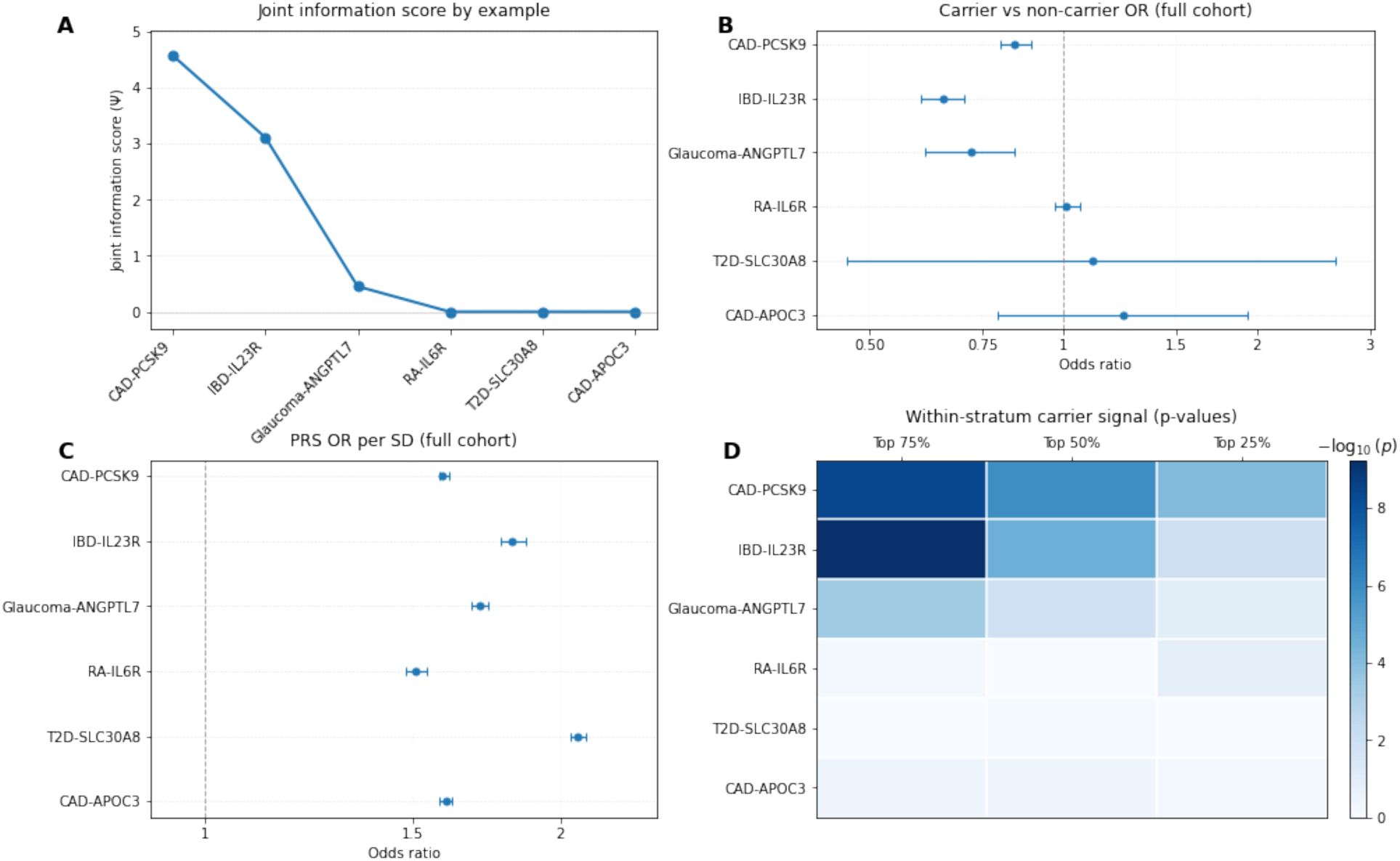
Evaluation of candidate disease–gene pairs for PRS-guided in silico enrichment. (A) Joint information score *Ψ* for each disease–gene example. (B) Carrier versus non-carrier odds ratios (ORs) with 95% confidence intervals estimated in the full cohort. (C) Odds ratio per standard deviation of PRS estimated in the full cohort. (D) Heatmap of within-stratum carrier association p-values across progressively enriched PRS strata (Top 75%, Top 50%, and Top 25%), shown as −*log*_10_(*p*).

These features are reflected in high joint information scores of 4.57 for CAD-PCSK9 and 3.11 for IBD-IL23R, supporting the use of these examples for evaluating PRS-guided prognostic enrichment within our framework.

Glaucoma-ANGPTL7 met baseline statistical criteria and showed meaningful stratification of disease risk by PRS. However, the difference in disease prevalence between carriers and non-carriers was attenuated and no longer statistically significant within the most restrictive Top 25% PRS stratum. Consistent with this pattern, the joint information score was lower at 0.45, reflecting more limited within-stratum information under aggressive restriction. We retained this example to illustrate how limited carrier–case observations under stringent enrichment thresholds can influence the performance of PRS-guided enrichment.

In contrast, RA-IL6R, T2D-SLC30A8, and CAD-APOC3 failed to demonstrate a statistically significant odds ratio comparing carriers with non-carriers in the full cohort and therefore did not meet the minimum criteria for enrichment evaluation in our dataset, although protective effects have been reported in previous studies. For RA-IL6R, the estimated carrier vs. non-carrier OR was 1.01 (95% CI: [0.97, 1.06]), consistent with the moderate effect size of the common variant p.Asp358Ala.^42,43^ For T2D-SLC30A8 and CAD-APOC3, the wide confidence intervals (T2D-SLC30A8: OR 1.11 [0.46–2.65]; CAD-APOC3: OR 1.24 [0.79–1.93]) likely reflect limited statistical power due to small numbers of variant carriers in the study population (66 carriers of *SLC30A8* p.Arg138Ter and 199 carriers of *APOC3* p.Ala43Thr). Correspondingly, the joint information scores of these examples were all zero, indicating insufficient signal for enrichment evaluation. These examples were therefore not pursued for further evaluation under PRS-guided prognostic enrichment.

### Effects of PRS and protective variant carrier status on disease prevalence

We analyzed 393,610 White British participants from the UK Biobank to evaluate the effects of protective genetic variants across PRS quantiles on disease prevalence for the three gene–disease pairs. The CAD–PCSK9 analysis included 41,697 CAD cases and 13,717 carriers of the *PCSK9* p.Arg46Leu variant. The glaucoma–ANGPTL7 analysis included 12,092 glaucoma cases and 6,314 carriers of the *ANGPTL7* p.Gln175His variant. The IBD-IL23R analysis included 6,172 IBD cases (2,436 Crohn’s disease and 4,396 ulcerative colitis) and 50,633 carriers of *IL23R* p.Arg381Gln.

Across all three disease contexts, carriers of the protective variants exhibited lower disease prevalence than non-carriers within each PRS stratum (Figure 3). In all examples, disease prevalence increased monotonically with higher PRS quantiles in both carriers and non-carriers, consistent with the expected positive association between polygenic risk and disease burden. At more stringent PRS thresholds, subgroup sample sizes reduced substantially, leading to larger uncertainty and reduced power to detect differences. For the glaucoma–ANGPTL7 example, given only 72 cases among 1,530 carriers in the Top 25% PRS stratum, the difference in disease prevalence between carriers and non-carriers was not statistically significant.

**Figure 3.**
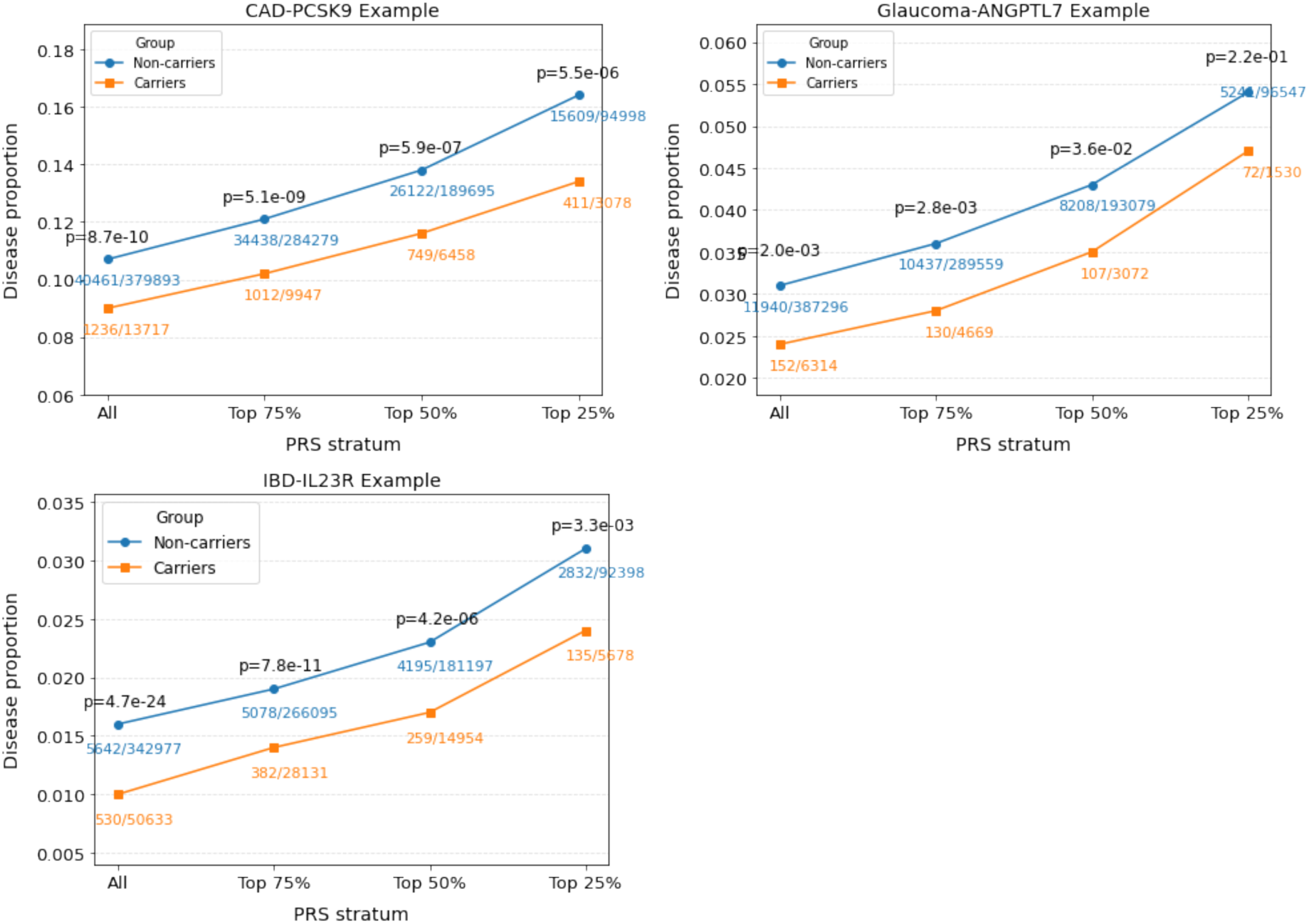
Disease prevalence in carriers versus non-carriers of protective variants across PRS strata for CAD–PCSK9, glaucoma–ANGPTL7, and IBD–IL23R. Numbers denote case counts and total sample sizes in each subgroup. P-values correspond to two-sided two-proportion tests comparing carriers and non-carriers within each stratum.

Logistic regression models adjusting for age, sex, and genetic principal components confirmed that carrier status was associated with lower disease risk across all three gene–disease pairs. For coronary artery disease, the *PCSK9* p.Arg46Leu variant was associated with an odds ratio of 0.878 (p = 0.003). For glaucoma, the *ANGPTL7* p.Gln175His variant was associated with an odds ratio of 0.760 (p = 0.004). For inflammatory bowel disease, the *IL23R* p.Arg381Gln variant was associated with an odds ratio of 0.913 (p = 0.049). In all three examples, PRS was strongly associated with disease risk (CAD PRS: OR = 1.438, p < 1 × 10^-16^; glaucoma PRS: OR = 1.843, p < 1 × 10^-16^; IBD PRS: OR = 1.85, p < 1 × 10^-16^). Complete model estimates are provided in Tables S3–S5, and model-predicted disease prevalence across PRS is shown in Figure S2.

### Sample size requirements and operational implications of PRS-enriched designs

Using model-based predictions of disease prevalence within each PRS stratum, we derived the minimum per-arm sample sizes required to achieve 60–85% statistical power under a two-sided 5% significance level across PRS-enriched subgroups (Figure 4). Across all three gene–disease pairs, restricting recruitment to higher PRS strata reduced required sample sizes. However, both the estimated per-arm sample size required in the unenriched population and the magnitude of reduction achieved under PRS enrichment differed substantially across disease contexts.

**Figure 4.**
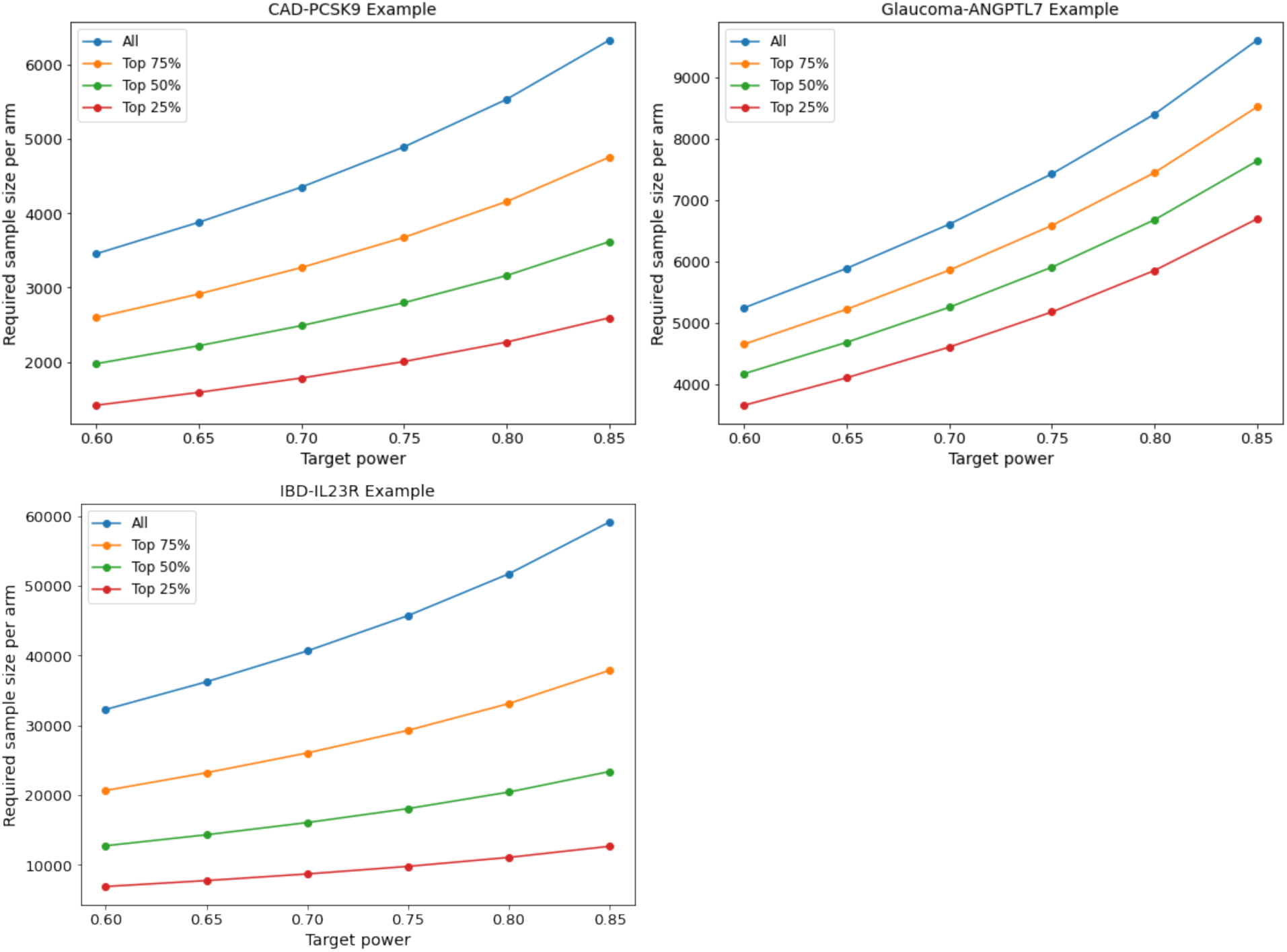
Estimated per-arm sample size required to achieve target power (60–85%) under PRS-guided enrichment, assuming 1:1 randomization and two-sided 5% significance level. Per-arm sample size requirements were derived from analytical power calculations based on disease prevalence predicted by logistic regression models for carriers and non-carriers of protective variants, adjusting for covariates as described in the Methods. Curves show the per-arm sample size required to achieve target power levels ranging from 60% to 85% across polygenic risk score (PRS) strata.

At 80% power, the CAD–PCSK9 example required 6,099 participants per arm in the full cohort. Restricting recruitment to the top 25% of the PRS distribution reduced this requirement to 2,383 per arm, representing an approximate 60% reduction. For glaucoma–ANGPTL7, the required per-arm sample size at 80% power was 8,404 in the full cohort and 5,853 in the Top 25% PRS group, corresponding to a more modest reduction of approximately 30%. In contrast, the IBD– IL23R example required substantially larger absolute sample sizes due to lower disease prevalence. At 80% power, 51,710 participants per arm were required in the full cohort, compared with 11,089 in the Top 25% PRS group. Despite the larger absolute numbers, this represented the greatest relative reduction (78%) among the three examples.

Projected total trial cost followed similar patterns (Figure S3). Under a general assumption that sequencing is required to obtain PRS, restriction to the Top 25% PRS group reduced projected total cost by 41.8% for CAD–PCSK9 and 69.5% for IBD–IL23R relative to the unenriched design. In glaucoma–ANGPTL7, however, the greatest relative cost reduction (9.3%) was observed at the Top 50% PRS threshold, whereas restriction to the Top 25% PRS group yielded only a 1.0% reduction. In contrast, under a scenario in which genetic data were assumed to be pre-existing and no additional sequencing cost was incurred, projected total trial cost decreased monotonically across PRS strata in all three examples. Restricting recruitment to the Top 25% PRS group reduced projected total cost by 51.7% for CAD–PCSK9, 17.8% for glaucoma– ANGPTL7, and 74.7% for IBD–IL23R.

### Empirical power under PRS-guided enrichment

We next evaluated empirical power through bootstrap simulations under fixed per-arm sample sizes of 100, 200, 500, and 1,000 participants (Figure 5). For the CAD–PCSK9 example, empirical power increased consistently with increasing PRS enrichment across all sample sizes, with the highest power consistently observed in the top 25% PRS group. At 1,000 participants per arm, empirical power increased from approximately 20% in the full cohort to 50% in the Top 25% PRS group.

**Figure 5.**
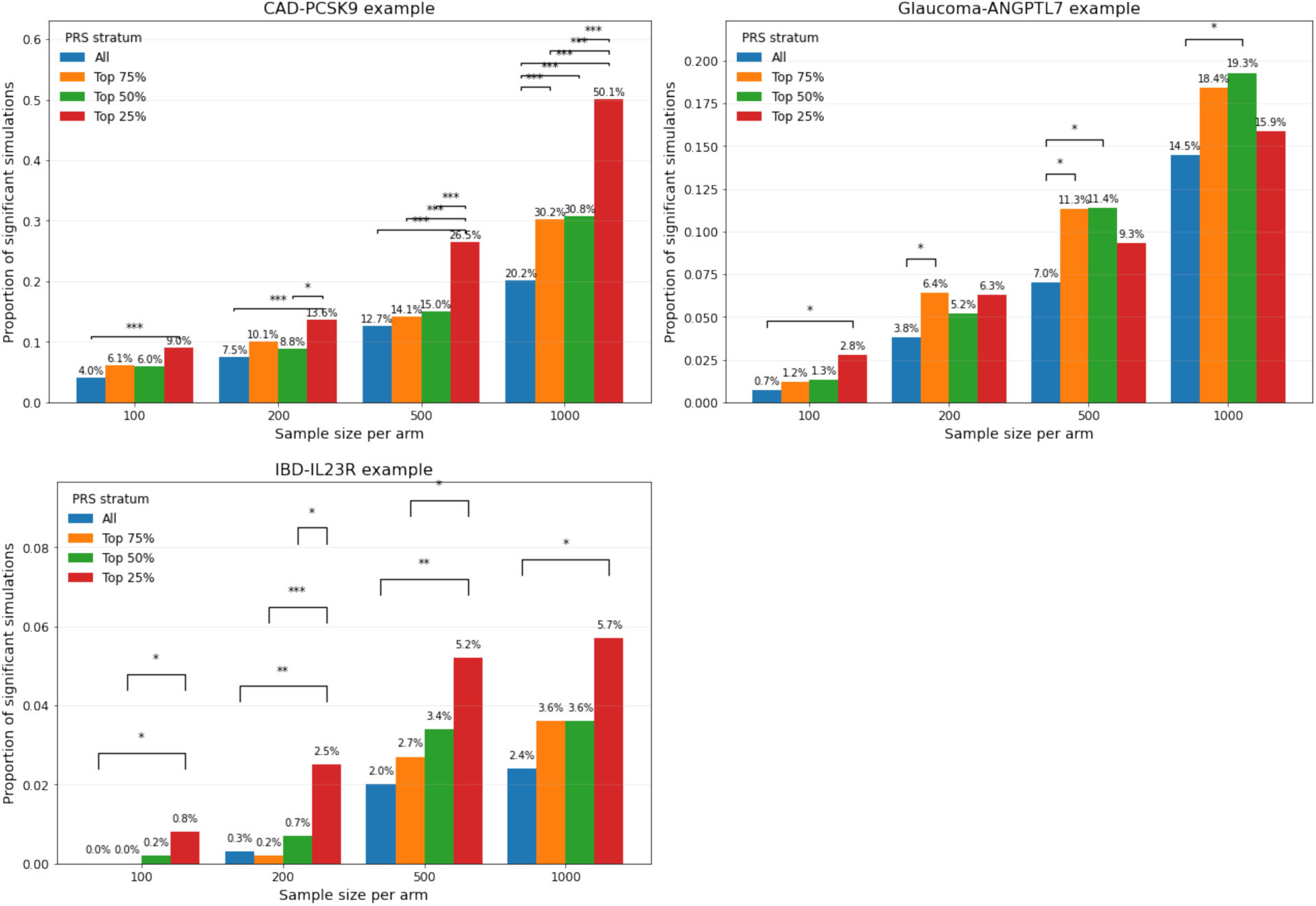
Empirical power of simulated trials with 100, 200, 500, or 1000 participants per arm. For each sample-size scenario, the proportion of significant simulations (empirical power) is compared across PRS strata. Pairwise differences between strata are tested using two-sided proportion tests, and significant contrasts after adjusting for multiple testing are annotated on the plot using standard significance codes (*** p < 0.001, ** p < 0.01, * p < 0.05).

A similar monotonic pattern was observed for the IBD–IL23R example, although absolute power remained modest even at larger sample sizes. At 1,000 participants per arm, empirical power increased from approximately 2–3% in the full cohort to about 5–6% in the Top 25% PRS group, reflecting the lower overall prevalence of IBD in the study population.

In contrast, the glaucoma–ANGPTL7 simulations did not show a consistent increase in empirical power across progressively higher PRS thresholds. While PRS-enriched groups generally outperformed the full cohort, the Top 25% PRS group did not uniformly achieve higher power than the Top 50% or Top 75% groups. At 1,000 participants per arm, empirical power increased from 14.5% in the full cohort to 18.4% and 19.3% in Top 75% and Top 50% PRS groups, respectively, but declined to 15.9% in the Top 25% PRS group. This non-monotonic pattern is consistent with the observation that disease prevalence did not differ significantly between carriers and non-carriers within the Top 25% PRS group due to substantially reduced number of carriers–case observations.

We note that in the IL23R–IBD and glaucoma–ANGPTL7 examples, empirical power fell below 5% in some small-sample scenarios, as our simulation classified comparisons between carriers and non-carriers as nonsignificant when fewer than five disease cases were observed in either resampled group.

### Prospective event accrual under PRS-enriched recruitment

We constructed baseline disease-free cohorts for the CAD–PCSK9, glaucoma–ANGPTL7, and IBD–IL23R gene–disease examples and followed participants for up to 10 years from UK Biobank enrollment. Baseline demographic characteristics and follow-up summaries are shown in Table 1. Age and sex distributions were similar across cohorts, whereas overall event counts differed substantially by disease context.

**Table 1.** Baseline characteristics of baseline disease-free cohorts for prospective event simulations. Demographic characteristics, follow-up duration, and total incident events for CAD–PCSK9, glaucoma– ANGPTL7, and IBD–IL23R cohorts. All participants were free of the relevant disease at baseline and followed for up to 10 years.

|  | CAD-PCSK9 | Glaucoma-ANGPTL7 | IBD-IL23R |
| --- | --- | --- | --- |
| N participants | 380678 | 392276 | 390820 |
| Age and baseline, mean (SD) | 56.7 (8.0) | 56.9 (8.0) | 56.9 (8.0) |
| Female, % | 44.9 | 45.9 | 45.9 |
| Number of events | 20122 | 9759 | 2485 |
| Median follow-up, years | 10.0 | 10.0 | 10.0 |

Across all three examples, distributions of the restricted mean survival time (RMST) consistently shifted downwards from the full cohort to the top 25% PRS group, indicating earlier accumulation of events within enriched groups (Figure 6). In the CAD–PCSK9 example, at 1,000 participants per arm, the mean RMST across 1,000 simulation replicates decreased from 9.77 years (95% CI: [9.71, 9.82]) in the unenriched cohort to 9.66 years (95% CI: [9.60, 9.73]) in the Top 25% PRS group. In the glaucoma–ANGPTL7 example, the mean RMST was 9.94 years (95% CI: [9.92, 9.97]) in the unenriched cohort and 9.88 years (95% CI: [9.85, 9.92]) in the Top 25% PRS group. In the IBD–IL23R example, the difference was more modest, with mean RMST of 9.97 years (95% CI: [9.95, 9.99]) in the unenriched cohort and 9.95 years (95% CI: [9.93, 9.98]) in the Top 25% PRS group.

**Figure 6.**
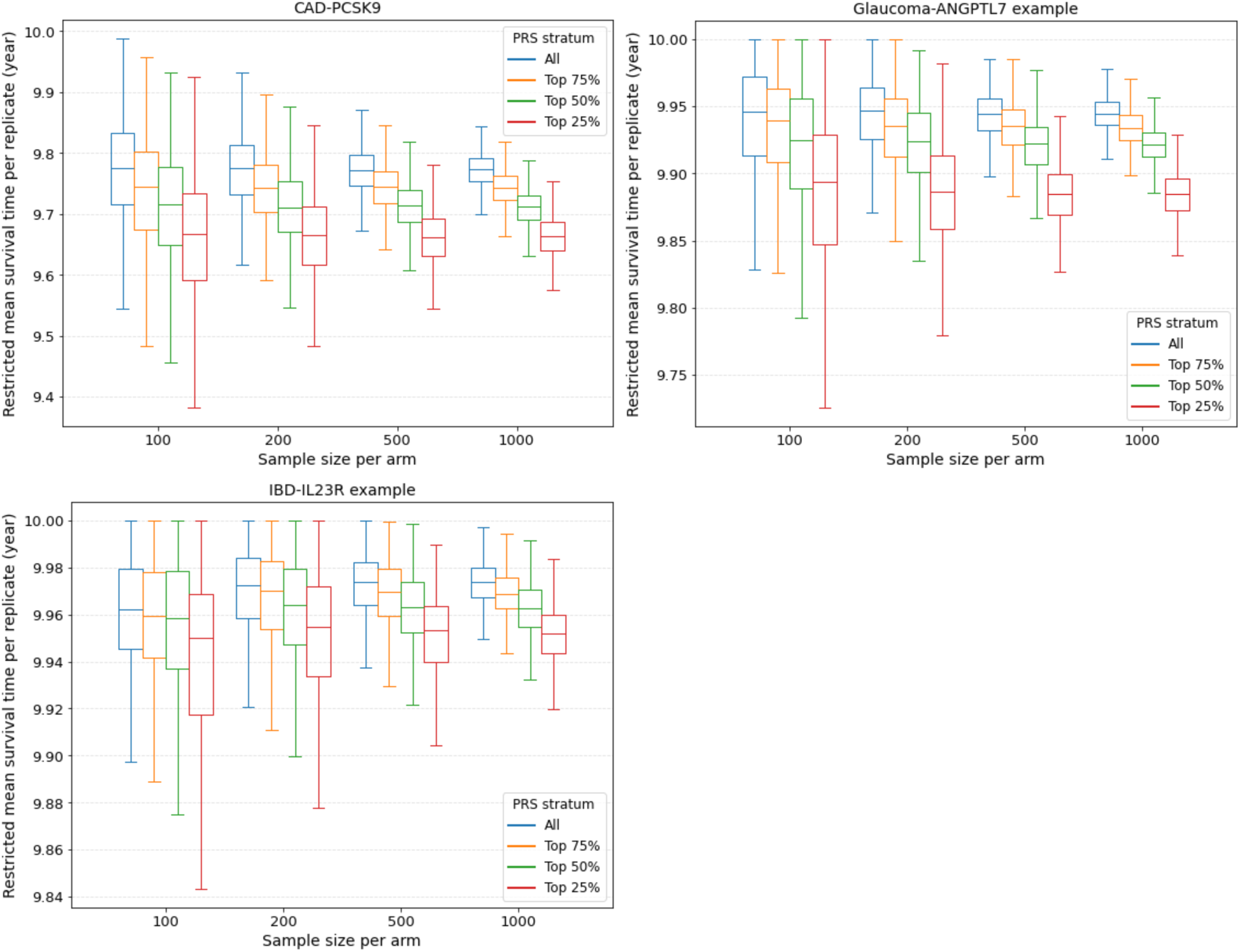
Restricted mean survival time under PRS-enriched recruitment. Distributions of restricted mean survival time (RMST) from fixed-size bootstrap simulations across PRS strata (All, Top 75%, Top 50%, Top 25%) and per-arm sample sizes (100, 200, 500, 1,000). RMST was calculated for up to 10 years of follow-up.

We note that RMST estimates depend on the chosen follow-up horizon and may also be sensitive to sampling variability. In our setting, the absolute differences in RMST were modest in part because the number of observed events was limited, particularly in the glaucoma–ANGPTL7 and IBD–IL23R examples. Variability in RMST estimates also increased in more restrictive PRS groups due to the reduced number of eligible participants and events within narrower PRS strata.

## Discussion

In this study, we present an in silico framework for prospectively evaluating how polygenic risk information can be incorporated into clinical trial design through prognostic enrichment using large-scale biobank data. By combining naturally occurring protective genetic variants as proxies for therapeutic interventions with PRS-based stratification of disease risk, we demonstrate how observed population data can be leveraged to inform key trial design parameters prior to trial initiation. We systematically reviewed disease–gene examples with established protective variant effects and assessed whether the available data provided sufficient statistical information to power the evaluation of PRS-guided enrichment in each setting. Our quantitative framework integrates population-level estimation of disease prevalence, analytical sample size calculation, simulation-based evaluation of empirical power and prospective event accrual, enabling systematic assessment of how PRS enrichment influences sample size requirements, statistical power, and the rate of event accumulation over follow-up.

Across three distinct disease systems illustrated by CAD–PCSK9, glaucoma–ANGPTL7, and IBD–IL23R use-cases, protective variant carriers consistently exhibited lower disease prevalence within PRS strata, and higher PRS was uniformly associated with increased disease risk. These shared features translated into reduced sample size requirements, lower projected operational costs, improved empirical power, and accelerated event accrual relative to unenriched designs. However, the magnitude and pattern of efficiency gains under PRS enrichment varied across disease contexts. In both the CAD-PCSK9 and IBD-IL23R example, the greatest enrichment benefits were observed in the most restrictive Top 25% PRS stratum. The glaucoma–ANGPTL7 example showed a similar pattern but more modest gains in sample size reduction and cost savings across progressively higher PRS strata, with moderate enrichment at the Top 50% threshold achieving empirical power and projected cost reductions comparable to, or exceeding, those obtained with the Top 25% threshold.

The variation in optimal PRS cutoffs across diseases highlights important operational considerations for implementing PRS-based enrichment in clinical trials. One key consideration is that the optimal enrichment threshold depends on the trade-off between enriching the cohort for higher disease risk and retaining a sufficiently large pool of eligible participants. In the CAD–PCSK9 and IBD–IL23R examples, enrichment to the upper PRS quartile was an efficient design choice, as it concentrated risk while preserving sufficient representation of protective variant carriers. In comparison, in the glaucoma–ANGPTL7 example, more moderate PRS thresholds provided comparable or superior efficiency by avoiding excessive loss of eligible participants. Another practical consideration is that increasingly stringent PRS thresholds require screening larger populations to identify eligible participants. When the cost of running the trial exceeds the cost of screening potential participants, as assumed in our model, reductions in required sample size generally outweigh the additional screening costs, especially when genetic data are already available. However, this trade-off depends on the relative costs of screening and trial execution, which vary across diseases and trial designs. In addition, more stringent enrichment criteria may require longer screening periods to identify eligible participants. Although our RMST analyses indicated a consistent pattern of earlier event occurrence in higher-risk cohorts, the absolute reductions in follow-up time were modest, and whether these gains would offset potentially longer recruitment periods would require further evaluation under realistic assumptions about trial recruitment and implementation. Together, our findings emphasize that PRS enrichment should be implemented through a structured, data-driven framework tailored to disease context, genetic risk distribution, and trial settings, rather than relying on a single uniform threshold.

Beyond methodological innovations, our work also presents broader implications for the clinical research and healthcare community. At present, germline genetic information is not routinely incorporated into clinical trials, despite the rapidly expanding availability of genomic data in population cohorts and healthcare systems. Our results contribute quantitative evidence demonstrating the potential value of human genetic data, including naturally occurring protective variants and polygenic risk scores, for informing clinical trial design. As such evidence accumulates, clinical trialists and sponsors may increasingly consider PRS-guided enrichment as a strategy to improve trial efficiency. In the meantime, governing bodies, trial registers, and adjudicating agencies may have increasing opportunities to develop guidance for genetically informed clinical trials. Moreover, while our study focuses on PRS and genetic data, the framework developed here is not limited to PRS-based enrichment. Other predictors, including molecular biomarkers, imaging measures, and clinical risk scores, may also serve as enrichment variables for clinical trials. The quantitative framework presented here provides a general approach for evaluating such enrichment strategies and may facilitate further comparison of alternative enrichment variables in future trial design studies.

While our framework provides a structured approach for quantitatively evaluating and optimizing PRS-guided enrichment in clinical trial design, several limitations warrant consideration. First, the implementation of this framework in the present study is constrained using a single population cohort, specifically the White British subset of the UK Biobank. Although UK Biobank is a well-characterized population-based resource, evaluation of enrichment performance is inevitably constrained by the available carrier counts and event numbers within the dataset, particularly for rare variants and low-prevalence diseases. In such settings, reduced statistical information may limit precision and render some gene–disease pairs underpowered for robust evaluation of enrichment effects. Integration of additional large-scale biobanks and meta-analyses across independent cohorts would increase carrier representation and event counts and enable more comprehensive evaluation of enrichment strategies, expanding application of this framework across a broader range of therapeutic targets and disease contexts. Such extension will be essential to further establish the generalizability and robustness of the proposed approach.

Second, PRS-guided enrichment raises considerations regarding the generalizability of trial results. Restricting enrollment to individuals with elevated PRS is intended to increase event rates and improve statistical efficiency, but treatment effects estimated in PRS-enriched populations may not directly generalize to individuals across the full PRS distribution. This limitation is an inherent challenge in trial enrichment strategies that restrict enrollment to a subgroup of the target population. A related consideration is potential labeling implications, as trials conducted in PRS-enriched populations may lead to indications tied to PRS-based eligibility. However, PRS can be viewed as one approach to identifying individuals at elevated disease risk, and the resulting evidence may be broadly informative for high-risk patients beyond those defined strictly by PRS, although the extent of this translation will depend on the clinical and regulatory context. In addition, our results derived from the UK Biobank White British cohort may also not directly generalize to populations with different genetic architectures, linkage disequilibrium patterns, or environmental exposures. In practice, enriched trials may provide an efficient approach for establishing initial evidence of therapeutic efficacy, while subsequent studies or post-marketing analyses would be necessary to evaluate effectiveness across broader patient populations.

Finally, our analyses rely on several assumptions related to the use of human genetic variation to inform therapeutic target evaluation. We assume that naturally occurring protective variants can serve as proxies for pharmacologic target modulation. Although this approach does not fully capture the complexity of therapeutic intervention, prior work has shown that human genetic perturbations often provide informative models of target inhibition and drug response, supporting the conceptual validity of this strategy.^53–55^ Nevertheless, because the protective effects evaluated here reflect naturally occurring genetic variation, they may underestimate the magnitude of effect achievable by a therapeutic intervention with stronger or more sustained target modulation. In addition, the effectiveness of PRS-guided enrichment depends on the accuracy and transferability of the PRS and the heritability of the target disease. For conditions with limited genetic contribution or poorly calibrated PRS, the benefits of enrichment may be attenuated.

In summary, our study demonstrates how population-scale genomic and health record data can be used to evaluate PRS-guided prognostic enrichment as part of early-stage trial planning. Rather than relying on retrospective analyses of completed trials, our results quantify how PRS-based enrichment is expected to influence event burden, statistical power, and operational considerations before a trial is conducted, while highlighting the difference of enrichment effects across disease contexts. These findings emphasize that the utility of PRS-guided enrichment is context dependent and should be assessed with respect to disease-specific prevalence, risk gradients, and feasibility considerations. Ultimately, this framework offers a scalable foundation for incorporating genomic risk information into the design of future clinical trials and may be extended to evaluate other prognostic enrichment variables using similar principles.

## Data Availability

The data used in this study are available from the UK Biobank resource upon application. Due to data access restrictions, the authors cannot share individual-level data.

## Author contributions

M.A.R. and J.W.OS. designed the study. R. C. led the analysis and manuscript writing. E.A., Y.L., J.W.OS., and L.T. contributed to the prognostic trial enrichment design approach. All authors contributed to the writing of the manuscript and figures generation. All authors reviewed the manuscript.

## Acknowledgements

This research has been conducted using the UK Biobank Resource under Application Number 24983 and 22282. Based on the information provided in Protocol 44532 the Stanford IRB has determined that the research does not involve human subjects as defined in 45 CFR 46.102(f) or 21 CFR 50.3(g). All participants in the UK Biobank provided written informed consent (more information is available at https://www.ukbiobank.ac.uk/2018/02/gdpr/). We thank all the participants in the UK Biobank study. Some of the computing for this project was performed on the Sherlock cluster. We would like to thank Stanford University and the Stanford Research Computing Center for providing computational resources and support that contributed to these research results.

## Supplementary Materials

### Supplementary Methods

#### Cost model for PRS-guided clinical trial enrichment

To quantify the operational implications of polygenic risk score (PRS)–based trial enrichment, we developed a simplified cost model that translates the PRS-specific sample size requirements into total projected trial cost. Our goal is to illustrate relative changes in projected cost across PRS thresholds rather than estimate absolute expenditures, which in practice depend strongly on disease-specific, assay-specific, and trial-specific factors. Accordingly, we characterize the relative change in projected cost when restricting enrollment to individuals above a given PRS threshold compared with an unenriched trial. Our cost model conceptualizes the trial workflow as three sequential cost components, including general screening, genotyping and PRS computation, and trial execution for randomized participants, each scaled according to the number of individuals required at that stage under a given PRS threshold.

For each PRS threshold *q*, defined as enrolling only individuals in the upper (1 − *q*) quantile of the PRS distribution, where the unenriched trial corresponds to *q* = 0, let *n*_*arm*_(*q*) denote the analytically derived per-arm sample size required to achieve a given target power. The total number of randomized participants is:

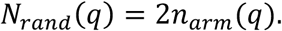

The number of individuals who must undergo genotyping and PRS assessment would then be

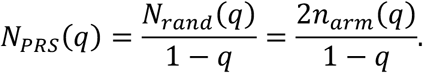

In practice, we might consider only genotyping individuals who pass the inclusion criteria and provide consent during an initial screening stage, during which a proportion *p*_*exclu*_ is excluded due to ineligibility or lack of consent. The total number of individuals requiring general screening is therefore

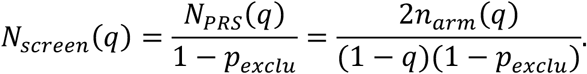

Suppose all individuals entering the screening incur a base screening cost *C*_*screen*_, and individuals who fail screening additionally incur a smaller incremental failure cost *C*_*fall*_. Then the cost during the screening stage would be:

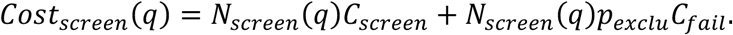

Assuming the cost of genotyping and PRS calculation is *C*_*geno*_, then the total cost to obtain PRS on individuals who passed the initial screening would be:

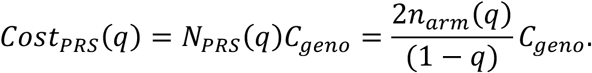

Finally, for participants whose PRS exceeding threshold *q*, assuming the cost of running the trial on each individual is *C*_*trial*_, which includes study visits, drug supply, safety monitoring, and endpoint adjudication, the total cost of trial execution would be:

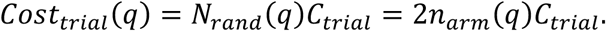

Combining the three components yields the total projected cost for a PRS-enriched design at threshold *q*:

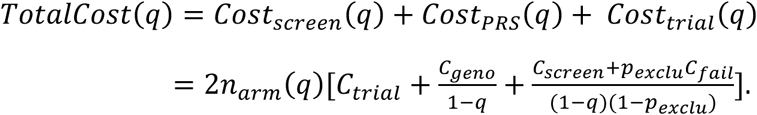

We define 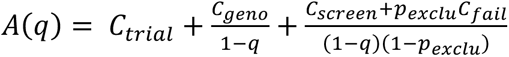 to represent the average per-randomized-participant cost at threshold *q*. The percent reduction in total cost of a PRS-enriched trial at threshold *q* relative to an unenriched trial can then be expressed as:

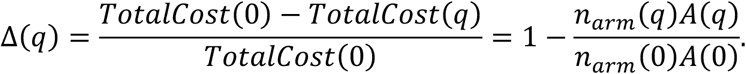

We produced illustrative numerical examples of Δ(*q*) using analytically derived sample size estimates for the CAD–PCSK9 and glaucoma–ANGPTL7 simulations, assuming *C*_*screen*_ = 500, *C*_*trial*_ = 10000, and *p*_*exclu*_=0.2. Recognizing that large population-genomic initiatives are making genomic data increasingly available, we evaluated two scenarios: one in which per-person sequencing costs are included (*C*_*geno*_ = 1000), and another assuming that genetic data are already available at scale and can be obtained at negligible marginal cost (*C*_*geno*_ = 0), as anticipated in future biobank-enabled health systems. Since *TotalCost*(*q*) is linear in *n*_*arm*_(*q*), percent changes in cost are invariant across power targets. Therefore, we evaluate cost reductions at a representative target of 80% power, which fully captures the behavior of the model. The result is shown in Figure S2.

## Supplementary Figures and Tables

**Figure S1.**
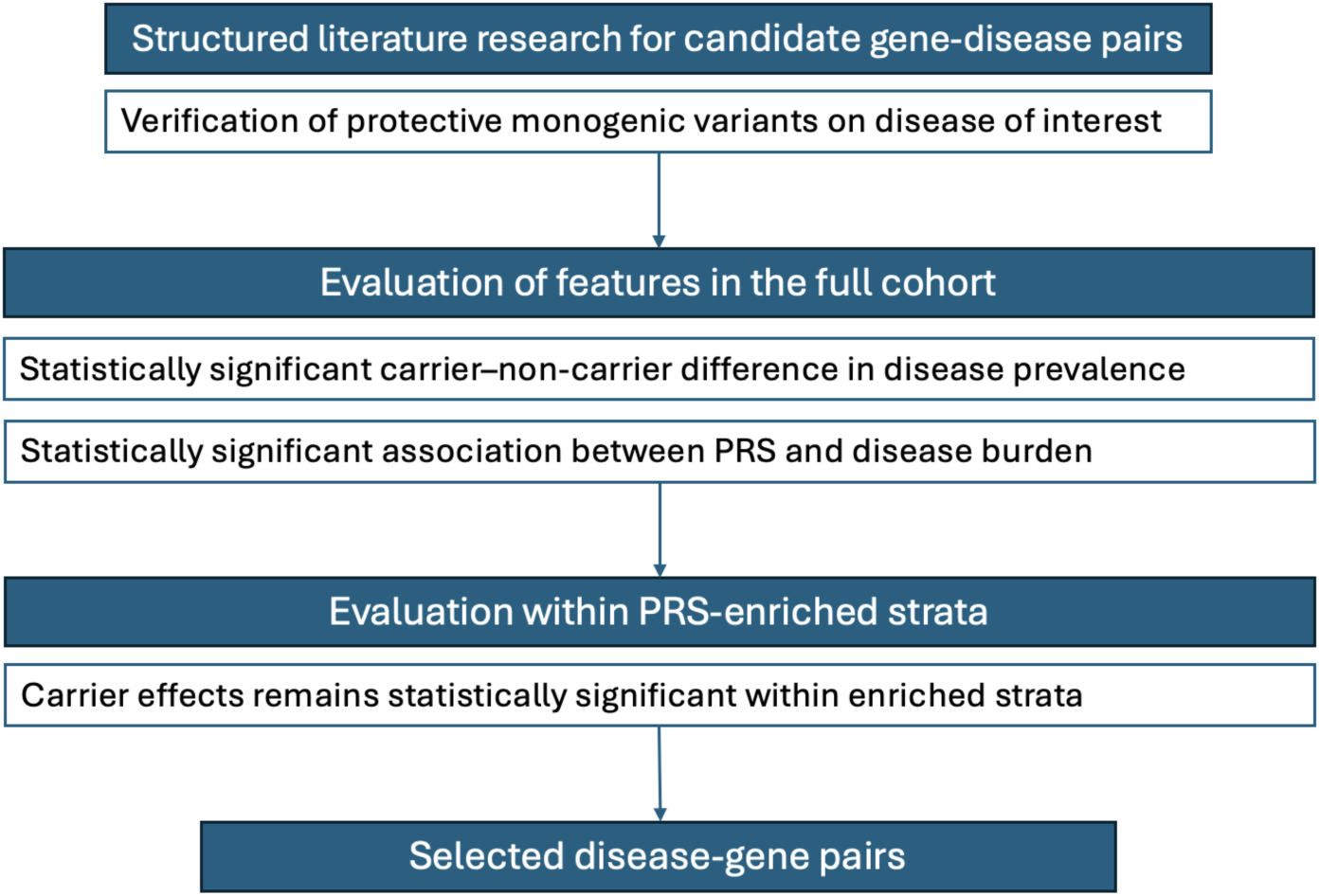
Workflow for selection of disease–gene pairs to evaluate PRS-guided in silico enrichment. Candidate disease–gene pairs involving biologically supported monogenic variants with established protective association to the disease of interest were identified through structured literature. These pairs were subsequently evaluated in the full cohort for a statistically supported carrier–non-carrier difference in disease risk and a significant association between polygenic risk score (PRS) and disease burden. Finally, we assessed whether the carrier–non-carrier contrast remained statistically supported within progressively enriched PRS strata. Disease–gene pairs satisfying these criteria were selected for full in silico enrichment analysis.

**Figure S2.**
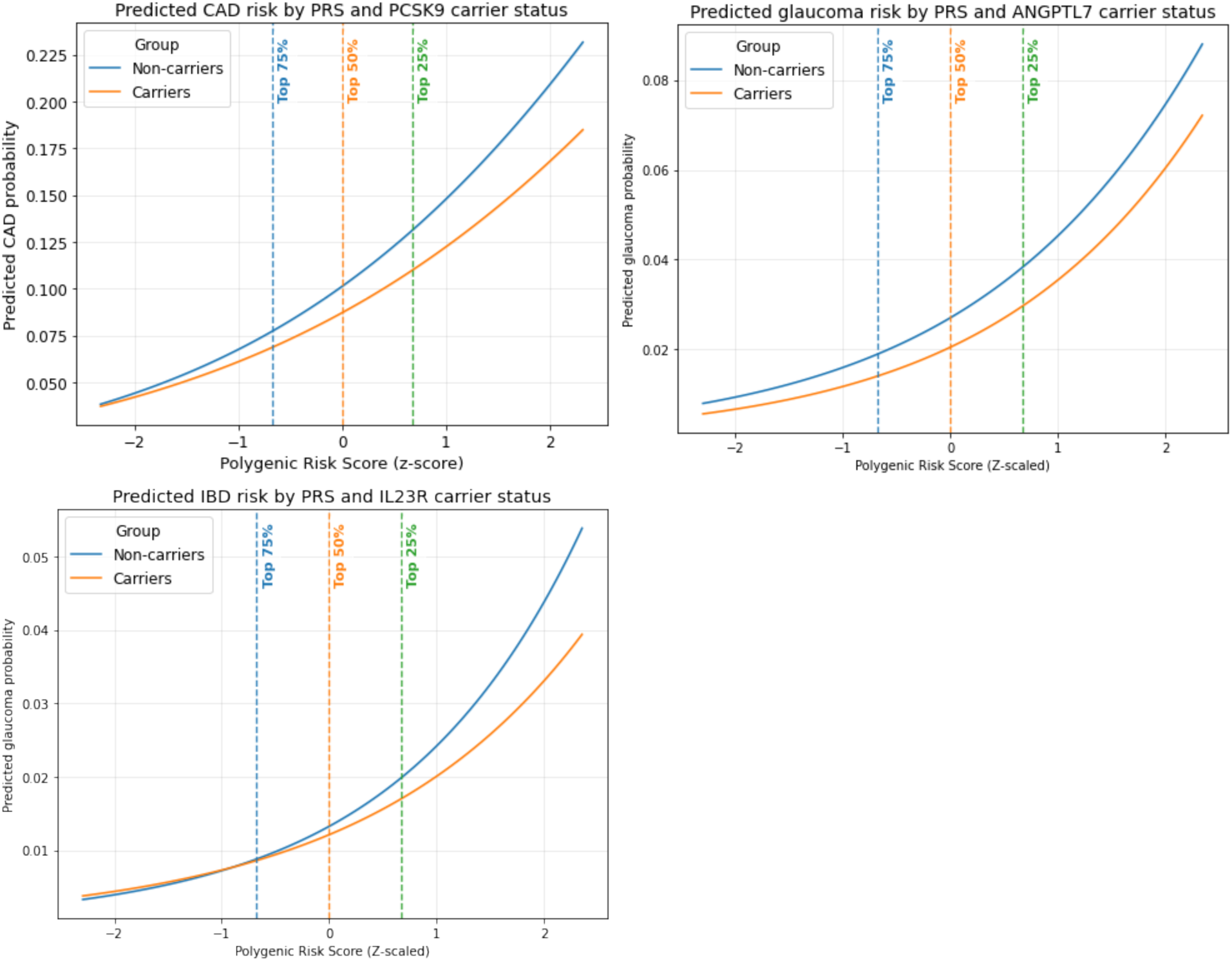
Predicted disease probabilities by logistic regression. Predicted disease risk was estimated using logistic regression models incorporating PRS, carrier status, and covariates. Curves represent the model-predicted risk for carriers and non-carriers across the continuous PRS distribution, with vertical dashed lines indicating PRS quantile thresholds used to define enrichment strata.

**Figure S3.**
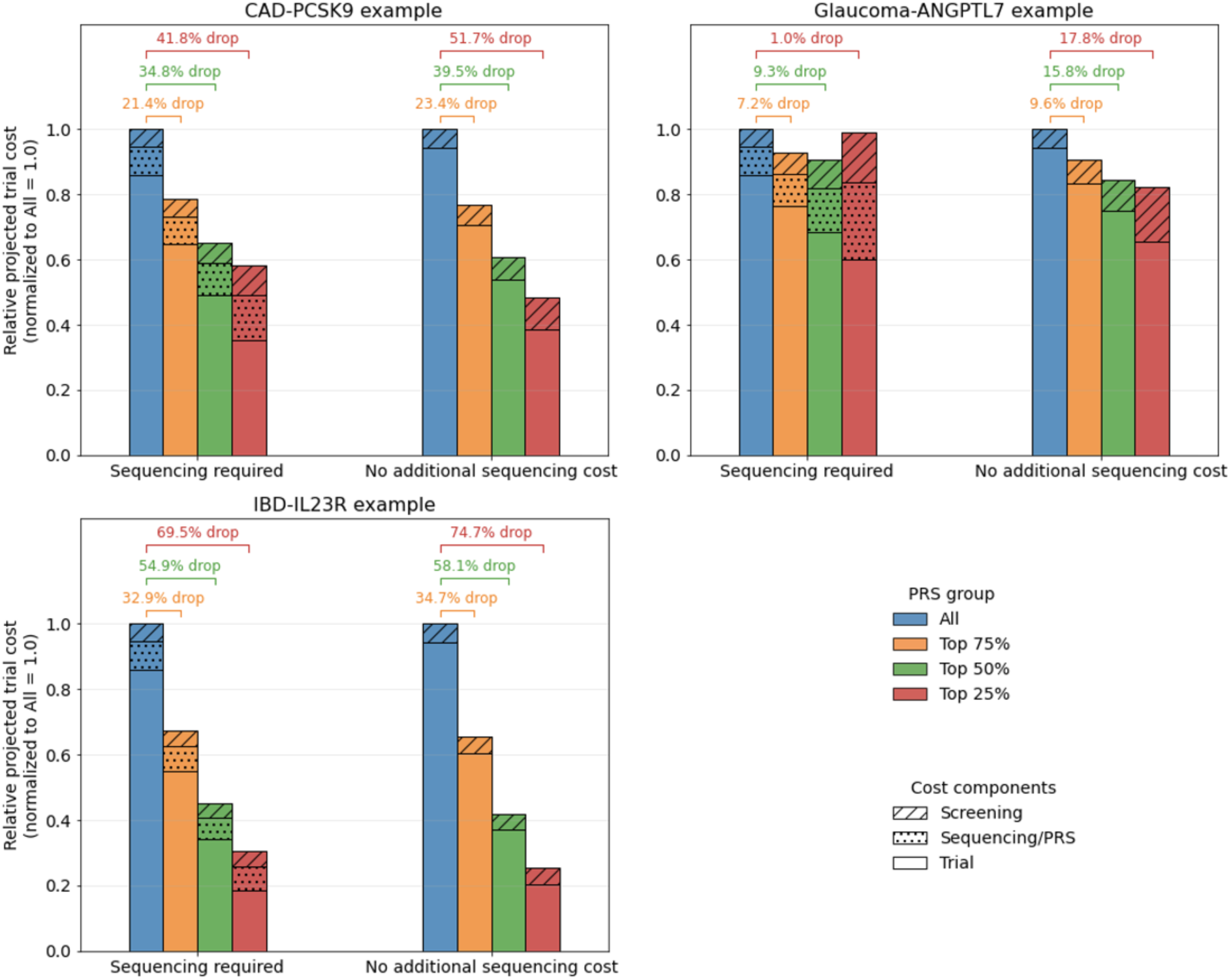
Relative projected trial cost under PRS-enriched recruitment. For each gene– disease example (CAD–PCSK9, glaucoma–ANGPTL7, and IBD–IL23R), bars display the total projected cost of a trial restricted to the top 75%, 50%, or 25% of the PRS distribution, scaled relative to an unenriched trial. The height of each bar corresponds to 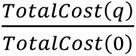. Annotated brackets indicate Δ(*q*), the percent reduction in overall cost compared with the full-population design. Bars are partitioned into cost components attributable to screening, sequencing/PRS, and per-participant trial costs, distinguished using different hatch patterns. Results are shown under two assumptions: a current-state scenario in which sequencing is required, and a future-state scenario in which no additional sequencing cost is incurred.

**Table S1.** Candidate monogenic variants and corresponding disease endpoint definitions. Disease–variant pairs identified through structured literature review and considered for evaluation under the PRS-guided enrichment framework. Variants are denoted using HGVS protein nomenclature. Disease endpoints were defined using ICD-10 codes as specified. Genomic coordinates correspond to the GRCh38 reference genome assembly.

| Variant | Genomic coordinates | Disease | ICD-10 codes for endpoint |
| --- | --- | --- | --- |
| *PCSK9* p.Arg46Leu | chr1:55039974 | CAD | I21 and I25, excluding I25.2 |
| *ANGPTL7* p.Gln175His | chr1:11252389 | Glaucoma | H40 |
| *IL23R* p.Arg381Gln | chr1:67705958 | IBD | K50, K51 |
| *IL6R* p.Asp358Ala | chr1:154426970 | RA | M05, M06 |
| *SLC30A8* p.Arg138Ter | chr8:118253964 | T2D | E11 |
| *APOC3* p.Ala43Thr | chr11:116701713 | CAD | I21 and I25, excluding I25.2 |

**Table S2.** Evaluation of disease–gene examples for implementing the PRS-guided in silico enrichment framework. Numbers of cases and participants in the full cohort and among variant carriers are reported. Baseline genetic and PRS signals are assessed in the full cohort using carrier versus non-carrier odds ratios and the odds ratio per standard deviation of PRS. Within-stratum signal is evaluated using the Wald statistic for the carrier contrast in progressively enriched PRS strata. OR: odds ratio. Z_carrier: Wald statistic from logistic regression of disease prevalence on carrier status within the specified PRS stratum.

|  | CAD<br>(PCSK9) | Glaucoma<br>(ANGPTL) | IBD<br>(IL23R) | RA<br>(IL6R) | T2D<br>(SLC30A8) | CAD<br>(APOC3) |
| --- | --- | --- | --- | --- | --- | --- |
| *Sample counts* |  |  |  |  |  |  |
| Full cohort (Cases/N) | $\frac{41,697}{393,610}$ | $\frac{12,092}{393,610}$ | $\frac{6,172}{393,610}$ | $\frac{8,254}{393,610}$ | $\frac{32,274}{393,610}$ | $\frac{41,697}{393,610}$ |
| Carriers (Cases/N) | $\frac{1,236}{13,717}$ | $\frac{152}{6,314}$ | $\frac{530}{50,633}$ | $\frac{5,353}{255,320}$ | $\frac{6}{66}$ | $\frac{25}{199}$ |
| *Genetic and PRS association in full cohort* |  |  |  |  |  |  |
| Carrier OR<br>(95% CI) | 0.84<br>(0.80, 0.89) | 0.72<br>(0.61, 0.84) | 0.65<br>(0.60, 0.70) | 1.01<br>(0.97, 1.06) | 1.11<br>(0.46, 2.65) | 1.24<br>(0.79, 1.93) |
| PRS OR per SD<br>(95% CI) | 1.59<br>(1.58, 1.61) | 1.71<br>(1.68, 1.74) | 1.82<br>(1.78, 1.87) | 1.51<br>(1.48, 1.54) | 2.07<br>(2.04, 2.10) | 1.60<br>(1.58, 1.62) |
| *Carrier effect within PRS strata* |  |  |  |  |  |  |
| Z_carrier (p-value),<br>Top 75% PRS | 5.86<br>(4.7e-09) | 3.52<br>(4.4e-04) | 6.20<br>(5.6e-10) | 0.51<br>(0.613) | 0.03<br>(0.974) | 0.85<br>(0.394) |
| Z_carrier (p-value),<br>Top 50% PRS | 4.85<br>(1.3e-06) | 2.49<br>(0.0129) | 4.23<br>(2.4e-05) | 0.19<br>(0.849) | 0.30<br>(0.763) | 0.83<br>(0.408) |
| Z_carrier (p-value),<br>Top 25% PRS | 3.94<br>(8.0e-05) | 1.61<br>(0.107) | 2.56<br>(0.0105) | 1.48<br>(0.139) | 0.02<br>(0.985) | 0.45<br>(0.651) |
| *Joint information score* |  |  |  |  |  |  |
| $\Psi$ | 4.57 | 0.453 | 3.112 | 0 | 0 | 0 |

**Table S3.** Logistic regression model of coronary artery disease on PRS and PCSK9 carrier status. Odds ratios (ORs) and 95% confidence intervals are estimated from a logistic regression model including carrier status of the *PCSK9* p.Arg46Leu variant, standardized polygenic risk score (PRS), their interaction term, and covariates (age, sex, and the first three genetic principal components). No statistically significant interactions were observed between PRS and protective variant carrier status.

|  | OR (95% CI) | p-value |
| --- | --- | --- |
| Intercept | 0.023 (0.020, 0.026) | < 0.0001 |
| Carrier | 0.878 (0.806, 0.956) | 0.003 |
| PRS | 1.438 (1.416, 1.460) | < 0.0001 |
| Carrier * PRS | 0.930 (0.854, 1.012) | 0.092 |
| Age | 1.083 (1.081, 1.086) | < 0.0001 |
| Sex (Male) | 2.321 (2.252, 2.393) | < 0.0001 |
| PC1 | 0.992 (0.975, 1.006) | 0.0004 |
| PC2 | 0.996 (0.983, 1.002) | 0.116 |
| PC3 | 1.001 (0.991, 1.010) | 0.880 |

**Table S4.** Logistic regression model of glaucoma on PRS and ANGPTL7 carrier status. Odds ratios (ORs) and 95% confidence intervals are estimated from a logistic regression model including carrier status of the *ANGPTL7* p.Gln175His variant, standardized glaucoma PRS, their interaction term, and covariates (age, sex, and the first three genetic principal components). No statistically significant interactions were observed between PRS and protective variant carrier status.

|  | OR (95% CI) | p-value |
| --- | --- | --- |
| Intercept | 0.020 (0.017, 0.023) | < 0.0001 |
| Carrier | 0.759 (0.631, 0.914) | 0.004 |
| PRS | 1.722 (1.690, 1.755) | < 0.0001 |
| Carrier * PRS | 1.028 (0.872, 1.211) | 0.741 |
| Age | 1.103 (1.100, 1.107) | < 0.0001 |
| Sex (Male) | 1.073 (1.034, 1.113) | 0.0002 |
| PC1 | 1.000 (0.988, 1.012) | 0.994 |
| PC2 | 1.005 (0.993, 1.018) | 0.402 |
| PC3 | 0.989 (0.977, 1.000) | 0.060 |

**Table S5.** Logistic regression model of IBD on PRS and IL23R carrier status. Odds ratios (ORs) and 95% confidence intervals are estimated from a logistic regression model including carrier status of the *IL23R* p.Arg381Gln variant, standardized glaucoma PRS, their interaction term, and covariates (age, sex, and the first three genetic principal components). A weak interaction effect was observed between PRS and protective variant carrier status.

|  | OR (95% CI) | p-value |
| --- | --- | --- |
| Intercept | 0.013 (0.010, 0.016) | < 0.0001 |
| Carrier | 0.912 (0.833, 0.999) | 0.049 |
| PRS | 1.843 (1.794, 1.893) | < 0.0001 |
| Carrier * PRS | 0.904 (0.826, 0.990) | 0.029 |
| Age | 1.010 (1.007, 1.013) | < 0.0001 |
| Sex (Male) | 1.141 (1.085, 1.200) | < 0.0001 |
| PC1 | 1.009 (0.993, 1.025) | 0.267 |
| PC2 | 1.010 (0.993, 1.027) | 0.268 |
| PC3 | 0.997 (0.980, 1.013) | 0.673 |

